# Cataract surgical burden and district level disparities in Bangladesh: A retrospective study

**DOI:** 10.64898/2026.08.15.26360490

**Authors:** Md. Saleh Ahmed, Moutushi Islam, Anthony Albert, Akhter Ferdoshe Jahan, Nusrat Lubna Islam, Tasruba Shahnaz, Chowdhury Mashrur Mahdee

## Abstract

**Objective:** To describe trends in cataract surgical volume, district-level surgical burden, and early postoperative visual outcomes among patients treated through a multi-district outreach programme in Bangladesh.

**Methods and Analysis:** This retrospective study was conducted using data from patients undergoing cataract surgery through the outreach eye-camp programme of Bashundhara Eye Hospital and Research Institute across eight districts of Bangladesh, between 2016 and 2025 (excluding 2021 because of COVID-19). Annual surgical volume trend was assessed using Poisson regression. Postoperative visual outcome on day 1 was categorized as good, borderline, or poor per WHO criteria. Univariable and multivariable ordinal logistic regression identified predictors of worse outcome.

**Results:** 1,929 cataract-surgery records were included. Surgical volume rose from 34 cases in 2016 to a peak of 677 in 2023 (IRR = 1.20; 95% CI: 1.180, 1.220; p-value< 0.001). Small incision cataract surgery (SICS) was used in 99.43% cases. On postoperative day 1, 70.09% of eyes had a good outcome, 19.44% borderline, and 10.47% poor. Increasing age was independently associated with worse outcome, with 2 to 3 times higher odds among patients over 70. District was independently associated with outcome, with Chapainawabganj and Kushtia having lower odds of worse outcome than Brahmanbaria. Sex was significant only in unadjusted analysis.

**Conclusion:** Surgical volume rose substantially over time. About seven in ten eyes achieved a good outcome on day 1, with age and district as the main predictors of worse outcome. Limitations include a single early assessment, exclusion of incomplete records, no standardized refraction, and unmeasured predictors.

**KEY MESSAGES:** *What is already known on this topic:* Cataract is the leading cause of avoidable blindness in Bangladesh and other low- and middle-income countries, but existing evidence on cataract surgical services in Bangladesh comes mainly from cross-sectional population surveys, which cannot track how surgical volume, district-level burden, or postoperative outcomes have changed over time.

*What this study adds:* Using ten years of registry data from a multi-district outreach programme, this study shows about 20% annual rise in cataract surgical volume, with near-exclusive use of small incision cataract surgery, and postoperative day-1 visual outcomes vary independently by patient age and district of residence.

*How this study might affect research, practice or policy:* These findings support continued scale-up of small incision cataract surgery, routine postoperative refraction to improve functional vision, and standardized outcome monitoring to guide more equitable allocation of eye-care resources in Bangladesh.

## INTRODUCTION

Cataract remains as the major cause of vision loss around the world. In 2020, 15 million people over the age of 50 were estimated to be blind and 79 million people over 50 were estimated to have moderate to severe cataract-related visual impairment globally [1]. Cataracts are the leading cause of preventable or treatable vision impairment worldwide, accounting for an estimated 94 million cases of distance vision impairment or blindness among the approximately 1 billion people living with vision impairment [2]. Globally, one in two individuals still lack access to cataract surgery when they need it [2]. Nearly 40% (17.0 million) of the 43.3 million blind people worldwide in 2020 were blind due to cataracts, with women disproportionately affected [3]. Low- and middle-income countries (LMICs) continue to bear a disproportionate share of this burden, with comparatively worse surgical outcomes than high-income countries [3].

An example of this trend is Bangladesh, a LMIC nation whose population is aging quickly. According to Bangladesh’s National Blindness and Low Vision Survey, cataracts are the most common cause of blindness in adults [4]. Surgical coverage is disproportionately higher for men, which increases the unmet burden for women [4]. The survey also found that policy responses focused on assigning surgically trained ophthalmologists to each district hospital, which shows the unequal distribution of surgical capacity in Bangladesh. According to a more recent national cross-sectional survey, among Bangladeshi individuals aged 40 and older, cataracts accounted for 84.3% of cases of impaired vision and blindness [5]. According to a district-level Rapid Assessment of Avoidable Blindness (RAAB) conducted in Satkhira district, cataract surgery coverage was relatively high (86.1%), and the annual surgical rate was only 547 per million people, which is insufficient to fulfill current needs [6]. However, a RAAB of Rohingya refugees in Cox’s Bazar revealed that cataract surgery coverage was 81.2% [7].

Facility-based, longitudinal data on how the volume of cataract surgeries has evolved over time, as well as how surgical burden and postoperative outcomes differ among different districts are missing from the current literature. The majority of the evidence currently available is derived from cross-sectional and population-based surveys, which are not intended to monitor changes in service delivery or compare postoperative outcomes over time.

Using retrospective data on cataract surgical services provided in eight districts in Bangladesh, the current study addresses this gap. The purpose of this study is to describe the trend in the number of cataract surgeries performed in Bangladesh, to identify differences in surgical volume at district level and to evaluate early postoperative visual outcomes and their predictors, in order to guide more equal distribution of resources for eye care.

## MATERIAL AND METHODS

### Study design, setting, and data collection

This was a retrospective, registry-based study of patients who underwent cataract surgery through the outreach eye-camp programme of Bashundhara Eye Hospital and Research Institute (BEHRI), a tertiary eye care facility based in Dhaka, Bangladesh. As part of its community outreach activities, BEHRI regularly conducts free eye camps at rural sites across multiple districts of Bangladesh. All surgical and camp-level records generated during these outreach activities between 2016 and 2025 were reviewed for this analysis. No camps were conducted in 2021 because of the COVID-19 pandemic, and that year was therefore excluded.

Camps were organized following prior discussions with local organizers and community partners to determine suitable sites and dates. About two weeks before each camp, community-level awareness activities and patient registration began. These activities included announcements via loudspeaker, leaflet distribution, poster displays, and announcements at local mosques to inform residents of the upcoming free eye camp. Interested individuals registered and were screened at the camp. Attendance was voluntary, with no charges at any stage. At each camp, a consultant ophthalmologist and a team of nurses and ophthalmic assistants travelled from BEHRI with diagnostic equipment to perform on-site screening, during which basic demographic and clinical information, including age, sex, and provisional diagnosis, was recorded for every patient screened.

Patients identified at the camp as having visually significant cataract and considered fit for surgery were referred to BEHRI in Dhaka for the operative stage of care, where they were transported and accommodated at no cost. A more detailed preoperative assessment was undertaken at this stage, with additional information recorded, including eye laterality, diagnosis, intraocular lens (IOL) power, fasting blood glucose, systolic and diastolic blood pressure.

Patients with known diabetes underwent blood glucose assessment, and surgery was performed only once glycemic control was confirmed to be adequate for safe operative and wound-healing outcomes. Informed consent was obtained from every patient prior to surgery.

Selected patients underwent a three-day hospital protocol. Preoperative evaluation and testing on Day 1, surgery on Day 2, and postoperative follow-up on Day 3, after which they were discharged and returned home. All procedures were performed at BEHRI by experienced surgeons, using either small incision cataract surgery (SICS) or phacoemulsification. A single postoperative review was conducted on postoperative day 1 for each patient, at which visual acuity was recorded using the Snellen chart.

All camp-screening and surgical data were entered and stored securely in BEHRI’s institutional database. For the purposes of this study, records from all camps conducted between 2016 and 2025 (2021 excluded) were retrospectively extracted. Of 2,570 total surgeries performed during this period, 2,330 were cataract surgeries. After excluding records with incomplete demographic, clinical, or outcome data, 1,929 records with complete information were included in the final analytic sample. This study was exempted from full ethical review by the Institutional Review Board of Bashundhara Eye Hospital and Research Institute, Dhaka, Bangladesh, as the study used routinely collected, de-identified data and involved no participant contact. Patients and the public were not involved in the design, conduct, reporting, or dissemination of this study, as it was a retrospective analysis of routinely collected clinical registry data.

### Statistical analysis

Descriptive statistics were used to summarize the demographic, clinical, and surgical characteristics of the study population. The trend in annual surgical volume was assessed using Poisson regression, with calendar year modeled as a continuous predictor and the trend expressed as an incidence rate ratio (IRR) with 95% confidence interval (CI). Chi-square tests and Fisher’s exact test were used to compare the distribution of demographic and clinical characteristics, across districts. Postoperative visual outcome (good (≥6/18), borderline (<6/18 to 6/60), or poor (<6/60))[8] was recorded as an ordinal three-level variable (1 = good, 2 = borderline, and 3 = poor). Univariable and multivariable ordinal logistic regression models were fitted to identify factors independently associated with visual outcome, with results expressed as unadjusted and adjusted odds ratios (OR) with 95% CI, reflecting the odds of a worse outcome category. A p-value <0.05 was considered statistically significant throughout the analysis. All analyses were conducted using Stata (version 17.0; StataCorp LP, College Station, Texas).

## RESULTS

Between 2016 and 2025 (excluding 2021), 2,570 surgeries were performed, of which 2,330 were cataract surgeries. Complete information was available from 1,929 records, which were included in the study. Table 1 shows the baseline characteristics of patients included in the study. Over half of the surgeries were performed on male patients (1,012; 52.46%), and the majority of patients were aged 60-69 years (778; 40.33%), followed by the 70-79 years (497; 25.76%). Surgery was performed more often on the right eye (1,084; 56.19%) compared to the left (845; 43.81%). Diabetes was present in 976 patients (50.60%) and hypertension in 678 patients (35.15%). By district, the largest share of patients was in Comilla (500; 25.92%) and Chapainawabganj (458; 23.74%), while Mymensingh accounted for the lowest (12; 0.62%).

**Table 1:** Baseline characteristics of patients undergoing cataract surgery in Bangladesh.

| Characteristics | n (%) |
| --- | --- |
| **Sex** |  |
| Male | 1012 (52.46) |
| Female | 917 (47.54) |
| **Age groups** |  |
| <50 | 100 (5.18) |
| 50-59 | 421 (21.82) |
| 60-69 | 778 (40.33) |
| 70-79 | 497 (25.76) |
| 80 and above | 133 (6.89) |
| **Eye** |  |
| Left eye | 845 (43.81) |
| Right eye | 1084 (56.19) |
| **District** |  |
| Brahmanbaria | 251 (13.01) |
| Chapainawabganj | 458 (23.74) |
| Comilla | 500 (25.92) |
| Kishoreganj | 212 (10.99) |
| Kushtia | 304 (15.76) |
| Madaripur | 51 (2.64) |
| Tangail | 141 (7.31) |
| Mymensingh | 12 (0.62) |
| **Hypertension** |  |
| No | 1251 (64.85) |
| Yes | 678 (35.15) |
| **Diabetes** |  |
| No | 953 (49.40) |
| Yes | 976 (50.60) |
| **Total** | **1929** |

Table 2 shows the annual distribution of cataract surgeries. Surgical volume increased over the study period, from 34 cases (1.76%) in 2016 to a peak of 677 cases (35.10%) in 2023. Poisson regression confirmed a significant upward trend (IRR = 1.20; 95% CI: 1.180, 1.220; p-value < 0.001), corresponding to an approximate 20% increase in surgical volume per year.

**Table 2:** Annual distribution of cataract surgeries from 2016 to 2025.

| **Year** | **Number of cataract surgeries, n (%)** |
| --- | --- |
| 2016 | 34 (1.76) |
| 2017 | 56 (2.90) |
| 2018 | 167 (8.66) |
| 2019 | 186 (9.64) |
| 2020 | 118 (6.12) |
| 2022 | 201 (10.42) |
| 2023 | 677 (35.10) |
| 2024 | 152 (7.88) |
| 2025 | 338 (17.52) |
| **Total** | **1929** |

Trend in annual surgical volume assessed by Poisson regression with year as a continuous predictor; incidence rate ratio (IRR) = 1.20 (95% CI: 1.180, 1.220), p = <0.001.

Table 3 illustrates that the most commonly used surgical technique was SICS, performed in 1,918 eyes (99.43%), while phacoemulsification was used in only 11 eyes (0.57%). A total of 1,352 eyes (70.09%) had a good postoperative visual outcome, while 375 (19.44%) had a borderline outcome and 202 (10.47%) had a poor outcome.

**Table 3:** Distribution of type of cataract surgery and postoperative visual outcomes.

| **Characteristics** | **n (%)** |
| --- | --- |
| **Type of cataract surgery** |  |
| SICS | 1918 (99.43) |
| Phacoemulsification | 11 (0.57) |
| **Postoperative visual outcome** |  |
| Good ($\geq 6/18$) | 1352 (70.09) |
| Borderline (<6/18 – 6/60) | 375 (19.44) |
| Poor (< 6/60) | 202 (10.47) |

Table 4 shows that the distribution of patients across districts differed by sex (p-value < 0.001), age group (p-value = 0.020), hypertension status (p-value < 0.001), and diabetes status (p-value < 0.001), indicating significant demographic and clinical heterogeneity in the study population by region.

**Table 4:**
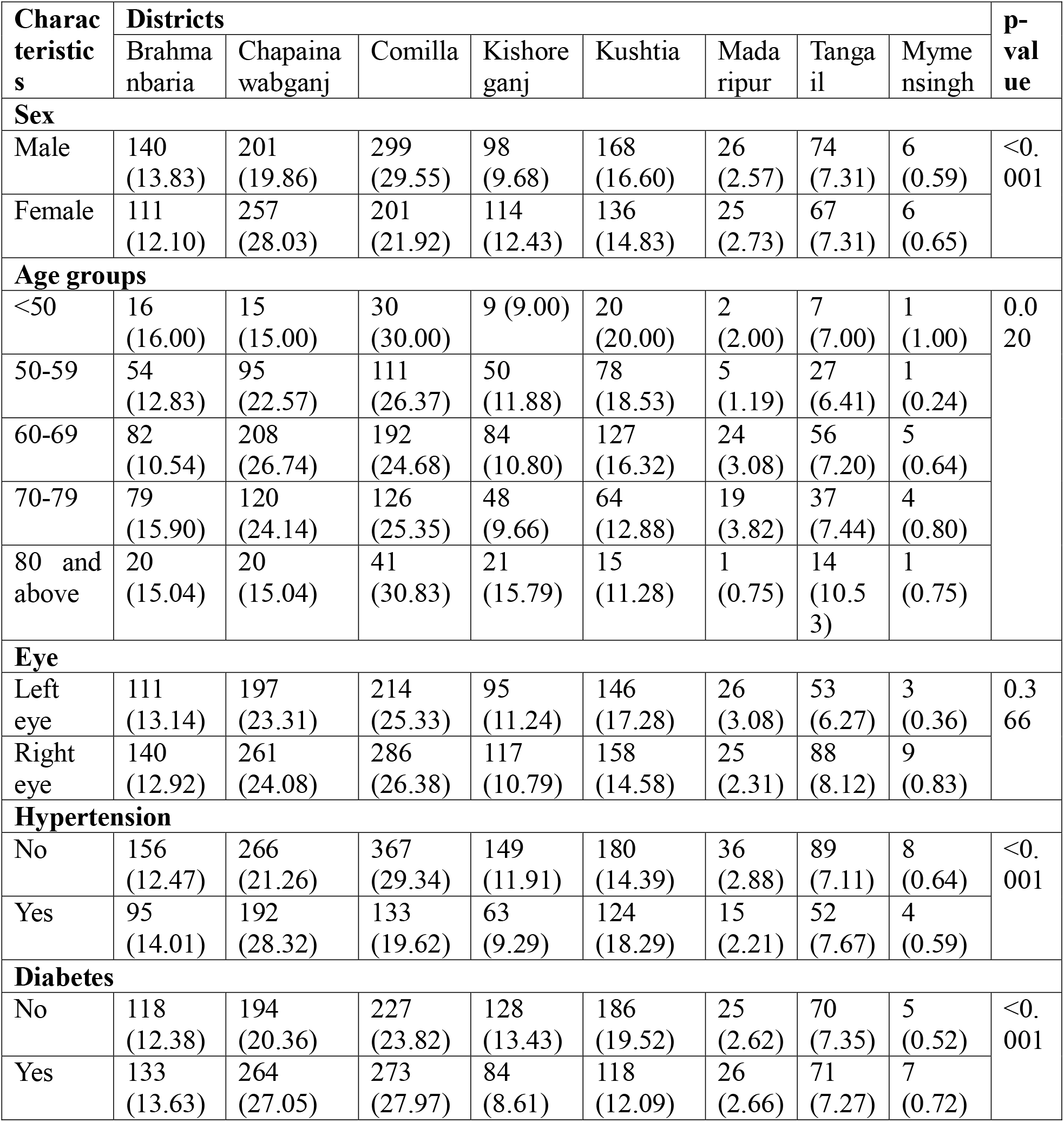
Comparison of demographic and clinical characteristics across districts.

Table 5 shows the associations between potential predictors and visual outcome following cataract surgery. Increasing age was significantly associated with worse visual outcome. Compared with patients younger than 50 years, those aged 70-79 years had more than twice the odds of a worse outcome (adjusted OR: 2.11; 95% CI: 1.26, 3.55; p-value = 0.005), and those aged 80 years and above had around three times the odds (adjusted OR: 3.06; 95% CI: 1.70, 5.53; p-value < 0.001). District was also independently associated with outcome, with patients from Chapainawabganj (adjusted OR: 0.65; 95% CI: 0.46, 0.91; p-value = 0.011) and Kushtia(adjusted OR: 0.61; 95% CI: 0.42, 0.88; p-value = 0.009) having significantly lower odds of a worse outcome compared to Brahmanbaria. Female sex was associated with lower odds of a worse outcome in the unadjusted model (unadjusted OR: 0.76; 95% CI: 0.63, 0.92; p-value = 0.005), but this association was diminished after adjustment (adjusted OR: 0.90; 95% CI: 0.73, 1.10; p-value = 0.303). Hypertension, diabetes, and eye laterality were not significantly associated with visual outcome in either the unadjusted or adjusted models. However, right eye surgery showed a borderline trend toward better outcome in the adjusted model (adjusted OR 0.82; 95% CI 0.68, 1.00; p-value = 0.051).

**Table 5:** Factors associated with postoperative visual outcome following cataract surgery.

| Factors | Unadjusted |  | Adjusted |  |
| --- | --- | --- | --- | --- |
|  | OR (95% CI) | p-value | OR (95% CI) | p-value |
| **Sex** |  |  |  |  |
| Male | Ref. | - | Ref. | - |
| Female | 0.76 (0.63, 0.92) | 0.005 | 0.90 (0.73, 1.10) | 0.303 |
| **Age groups** |  |  |  |  |
| <50 | Ref. | - | Ref. | - |
| 50-59 | 1.07 (0.63, 1.81) | 0.812 | 1.09 (0.64, 1.85) | 0.760 |
| 60-69 | 1.43 (0.86, 2.36) | 0.166 | 1.49 (0.89, 2.47) | 0.128 |
| 70-79 | 2.15 (1.29, 3.58) | 0.003 | 2.11 (1.26, 3.55) | 0.005 |
| 80 and above | 3.26 (1.83, 5.83) | <0.001 | 3.06 (1.70, 5.53) | <0.001 |
| **District** |  |  |  |  |
| Brahmanbaria | Ref. | - | Ref. | - |
| Chapainawabganj | 0.62 (0.45, 0.87) | 0.005 | 0.65 (0.46, 0.91) | 0.011 |
| Comilla | 0.99 (0.73, 1.36) | 0.989 | 1.03 (0.75, 1.42) | 0.853 |
| Kishoreganj | 0.83 (0.57, 1.22) | 0.352 | 0.85 (0.57, 1.26) | 0.413 |
| Kushtia | 0.57 (0.40, 0.83) | 0.003 | 0.61 (0.42, 0.88) | 0.009 |
| Madaripur | 0.99 (0.53, 1.87) | 0.992 | 0.99 (0.53, 1.87) | 0.985 |
| Tangail | 1.37 (0.90, 2.07) | 0.142 | 1.37 (0.90, 2.08) | 0.147 |
| Mymensingh | 0.45 (0.10, 2.12) | 0.313 | 0.43 (0.09, 2.06) | 0.289 |
| **Eye** |  |  |  |  |
| Left eye | Ref. | - | Ref. | - |
| Right eye | 0.84 (0.69, 1.02) | 0.076 | 0.82 (0.68, 1.00) | 0.051 |
| **Hypertension** |  |  |  |  |
| No | Ref. | - | Ref. | - |
| Yes | 1.01 (0.82, 1.23) | 0.961 | 1.03 (0.84, 1.27) | 0.783 |
| **Diabetes** |  |  |  |  |
| No | Ref. | - | Ref. | - |
| Yes | 0.96 (0.79, 1.17) | 0.703 | 0.94 (0.77, 1.15) | 0.564 |

## DISCUSSION

This retrospective analysis of 1,929 cataract surgeries across eight districts of Bangladesh (2016-2025) shows a significant rise in surgical volume over time, near-exclusive reliance on SICS, and visual outcomes that are favorable but below WHO benchmarks, with age, district, and sex (in unadjusted analysis) being associated with poorer outcomes. The approximate 20% annual rise in surgical volume (IRR = 1.20) parallels what is seen at other LMIC eye-care networks, such as the sustained growth documented at Aravind Eye Hospitals over a comparable period [9]. The decline in 2020 and the absence of recorded surgeries in 2021 reflect COVID-19-related disruption to elective surgical and outreach services.

The near-total dominance of SICS (99.4%) over phacoemulsification (0.6%) is more pronounced than at L V Prasad Eye Institute’s rural secondary centres (91.8% SICS) [10]. This fits the rationale for SICS in resource-limited settings as multiple studies found that SICS achieves outcomes comparable to phacoemulsification while being faster and less equipment-dependent [11–13]. But this contrasts with the gradual shift toward phacoemulsification seen at larger, better-resourced Indian centres [9], suggesting that further scale-up of SICS, rather than early investment in phacoemulsification, may be the more equitable near-term strategy for Bangladesh’s district-level services. Visual outcomes found in this study (70.1% good, 19.4% borderline, 10.5% poor) fall short of the WHO-recommended thresholds for presenting visual acuity [8]. These targets are formally defined for evaluation at four or more weeks after surgery, when transient postoperative inflammation, corneal edema, and induced astigmatism have resolved. WHO guidance for outcomes assessed at discharge or on postoperative day 1 instead sets a more lenient benchmark of over 50% good and under 10% poor outcomes [14]. Measured against this threshold, the good-outcome rate observed in this study exceeds the recommended rate, and the poor-outcome rate is very close to the recommendation, suggesting that a substantial part of the shortfall against the long-term WHO standard reflects the very early timing of assessment. These findings are comparable to the outcomes reported in multiple studies conducted in India and China [10,15–17], and are better than a population-based study in Satkhira, Bangladesh [18]. The best-corrected outcomes in Indian rural secondary centres rose sharply to 91.7% good once refractive correction was applied at 4-11 weeks [10], indicating that uncorrected refractive error is likely the dominant driver of suboptimal presenting-acuity outcomes in this study. This underscores the importance of routine post-operative refraction and affordable spectacle provision as a low-cost lever to substantially improve functional visual outcomes in the study population.

Increasing age was independently associated with worse outcome, with two to three times higher odds in patients aged 70-79 and ≥80 respectively, which is similar to findings from similar studies [10,19]. In rural Indian secondary centres, patients aged 70 years and older had markedly higher odds of a poor outcome [10], and at Jimma University Medical Center, a comparable age group had a similarly high odds [19]. District was also independently associated with outcome, with Chapainawabganj and Kushtia showing significantly lower odds of poor outcome than Brahmanbaria. Patients travelling from districts with more limited healthcare infrastructure or greater geographic isolation may face steeper barriers to reaching screening camps, leading them to present later with denser cataracts or undiagnosed ocular co-morbidity. These advanced preoperative states can increase surgical complexity and prolong early recovery, worsening the recorded day-1 outcome independent of the underlying surgical technique. This pattern is consistent with other Bangladeshi data linking geographic and socioeconomic disparities to cataract surgical outcome and service uptake more broadly [4,6,7]. This also suggests that at least part of the district-level variation observed in this registry can be described by patient-side barriers to timely care. Female sex was associated with worse outcome in unadjusted analysis. However, this association was lost after adjustment, diverging from similar studies, where the association persisted after adjustment [10], but aligning with Bangladesh’s own national survey, which found no independent sex difference in outcome [20]. Given that national surveys consistently show lower surgical coverage among women [4], inequity in this setting may lie more in access to surgery than in outcomes among those who receive it. Hypertension, diabetes, and eye laterality showed no significant association with outcome. This is consistent with evidence that diabetes without retinopathy does not meaningfully worsen outcomes, and that retinopathy severity is driving most of the excess risk in diabetic cataract populations [21,22].

Several limitations of this study should be considered. First, visual outcome was assessed at a single postoperative day-1 review, with no later clinical follow-up. This very early timepoint captures transient corneal edema and inflammation that inherently understate the visual acuity. Second, this was a retrospective registry analysis and 401 of the 2,330 (17.2%) cataract-surgery records were excluded because of incomplete demographic, clinical, or outcome data. So, the direction and magnitude of any resulting selection bias could not be assessed. Third, the registry did not capture several established predictors of cataract surgical outcome, including preoperative ocular co-morbidity, intraoperative complications, and surgeon experience, all of which have been shown to significantly influence outcome in comparable studies [10,19]. Fourth, outcome was not based on a standardized postoperative refraction, so some borderline and poor outcomes may reflect uncorrected refractive error rather than a limit on visual potential. Fifth, because surgical volume was derived from routinely collected administrative records rather than a fixed reporting framework, part of the observed rise over time may be due to improvements in data capture and camp coverage instead of a purely proportional increase in underlying surgical activity. Finally, as a multi-district but registry-based sample, these findings may not generalize to patients who never presented for surgery. Despite these limitations, this study offers one of the largest and most recent facility-based assessments of cataract surgical volume, technique, and early visual outcome across multiple districts of Bangladesh, addressing a previously identified gap in facility-based, longitudinal data on cataract surgical burden and outcome in this setting [4,6,7,18,20].

These findings point to two actionable priorities for district-level services in Bangladesh; routine post-operative refraction and spectacle provision to close the gap between surgical success and functional vision, and closer scrutiny of district-level variation in outcome to identify which structural or process factors underlie the better performance seen in Chapainawabganj and Kushtia relative to Brahmanbaria. As surgical volume continues to rise nationally, embedding standardized outcome monitoring, such as the WHO cataract surgical record used in comparable South Asian settings [10,18], would allow these district-level differences to be tracked and addressed prospectively rather than retrospectively. In conclusion, this multi-district registry-based study found a substantial rise in cataract surgical volume in Bangladesh between 2016 and 2025, driven predominantly by SICS. Approximately seven in ten eyes achieved a good visual outcome on postoperative day 1, with age and district of residence as the main independent predictors of a worse outcome. These findings support continued expansion of surgical capacity alongside routine outcome monitoring to guide quality improvement at the district level.

## Data Availability

All data produced in the present study are available upon reasonable request to the authors

## ACKNOWLEDGEMENT

The authors thank the staff of BEHRI involved in the outreach eye-camp programme for their contribution. This study was funded internally through the Zakat donation scheme of BEHRI.

## COMPETEING INTEREST

The authors declare no conflict of interest.

